# Genetics of Height and Risk of Atrial Fibrillation: A Mendelian Randomization Study

**DOI:** 10.1101/2019.12.23.19015743

**Authors:** Michael G. Levin, Renae Judy, Dipender Gill, Marijana Vujkovic, Regeneron Genetics Center, Matthew C. Hyman, Saman Nazarian, Daniel J. Rader, Benjamin F. Voight, Scott M. Damrauer

## Abstract

**Objective:** To determine whether height has a causal effect on risk of atrial fibrillation

**Design:** Mendelian randomization study

**Setting:** Genome-wide association studies of height and atrial fibrillation; Penn Medicine Biobank

**Participants:** Multiethnic (predominantly European ancestry) participants in genome-wide association studies of height (693,529 individuals) and atrial fibrillation (65,446 cases and 522,744 controls); 7,023 Penn Medicine Biobank participants of European ancestry

**Exposures:** Height, cardiometabolic risk factors for atrial fibrillation, and randomly allocated genetic variants strongly associated with these traits

**Main outcome measure:** Risk of atrial fibrillation (measured in odds ratio)

**Results:** At the population level, a 1 standard deviation increase in genetically-predicted height was associated with increased odds of AF (Odds ratio [OR] 1.34; 95% Confidence Interval [CI] 1.29 to 1.40; p = 5×10^−42^). These findings remained consistent in sensitivity analyses that were robust to the presence of pleiotropic variants. Results from analyses considering individual-participant data were similar, even after adjustment for clinical covariates, including left atrial size.

**Conclusions:** Genetically predicted height is a positive causal risk factor for AF. This finding raises the possibility of investigating height/growth-related pathways as a means for gaining novel mechanistic insights to atrial fibrillation, as well as incorporating height into population screening strategies for atrial fibrillation.

## INTRODUCTION

Atrial fibrillation is a common cardiac arrhythmia, with a population prevalence of 0.5%, affecting more than 33 million individuals worldwide.[1] A number of risk factors are associated with atrial fibrillation, including chronic diseases like chronic kidney disease, heart failure, hyperthyroidism, obesity, obstructive sleep apnea, sleep apnea, and valvular heart disease, as well as cardiac surgery, smoking, and anthropometric factors.[2–5] Recent studies have identified both common and rare genetic variants at more than 100 independent loci contributing to the incidence of atrial fibrillation, and heritability is estimated at 20%.[6–8] Even with treatment, affected individuals are at risk of cardioembolic stroke, heart failure, and death.[2]

Height has been identified as a risk factor for a number of cardiometabolic diseases, including coronary artery disease, atrial fibrillation, and venous thromboembolism.[9,10] The relationship between height and atrial fibrillation in particular has been identified in large observational studies, with greater height strongly associated with increased risk of atrial fibrillation.[4,11–18] These studies are limited in assessing the causality and potential mediators of confounders of this association by their observational design, and randomized controlled trials of anthropometric traits are not feasible. Although a number of factors influence height including childhood illness/development, diet/nutrition, and socioeconomic status, height has a strong genetic component. Large genome-wide association studies have provided heritability estimates of 60-70%, and identified more than 700 independent loci that contribute to height.[19–22]

In the current study, we utilize human genetic data within the Mendelian randomization framework to evaluate a potential causal association between height and atrial fibrillation. Mendelian randomization leverages the random assortment of genetic variants during meiosis as instrumental variables to estimate the causal role between a trait and an outcome of interest. Here, we use summary data from large multiethnic GWAS of height (N ∼ 693,529 individuals) and atrial fibrillation (65,446 cases and 522,744 controls) to estimate the effect of genetically-predicted height on risk of atrial fibrillation.[7,19] We then use participant level data from the Penn Medicine Biobank within the observational phenome-wide association study (PheWAS) framework to identify other clinical associations with height, and within the single-sample MR framework to further assess the impact of height on atrial fibrillation after adjustment for clinical risk factors.

## MATERIALS AND METHODS

### Mendelian Randomization Analysis

Summary-level data for GWAS of height and atrial fibrillation were obtained. Two-sample mendelian randomization, using summary-level data, was performed using the *TwoSampleMR* package in R.[23] Using variants associated with height at genome-wide significance (p < 5×10^−8^), SNPs were harmonized with the variants from the Roselli et al. (2018) atrial fibrillation GWAS, and LD-pruned (distance threshold = 10,000kb, r^2^ = 0.001) using the *clump_data* command in the *TwoSampleMR* package in R, identifying a final set of 707 independent SNPs to use as a genetic instrument for height, which accounted for 11.2% of the variability in height **(Supplemental Table 1)**. Inverse variance weighted two-sample Mendelian randomization was used as the primary analysis, with weighted median, MR-Egger, and MR-PRESSO performed as sensitivity analyses to account for potential violations of the instrumental variable assumptions.[24] Further sensitivity analysis was performed using a genetic instrument for height constructed excluding any SNPs nominally (p < 0.05) associated with coronary artery disease, HDL, LDL, total cholesterol, triglycerides, fasting glucose, fasting insulin, diabetes, BMI, waist-to-hip ratio, and systolic blood pressure, all identified using the MR-base database.[25–28]

For each variant included in the genetic instruments, variance (R^2^) was calculated using the formula *R*^2^ = 2 × *MAF* × (1 − *MAF*) × *beta*^2^ (where MAF represents the effect allele-frequency and beta represents the effect estimate of the genetic variant in the exposure GWAS). F-statistics were then calculated for each variant using the formula 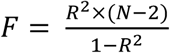 (where R^2^ represented the variance in exposure explained by the genetic variant, and N represents the number of individuals in the exposure GWAS) to assess the strength of the selected instruments.[29]

To account for the possibility that the restrictive genetic instrument may introduce collider or ascertainment bias by conditioning on associated traits, multivariable MR was performed using the *TwoSampleMR* package in R. This method allows the direct effects of multiple traits on an outcome to be determined jointly. The effect of height on atrial fibrillation was estimated in analyses that individually adjusted for potential confounders including coronary artery disease, HDL, LDL, total cholesterol, triglycerides, fasting glucose, fasting insulin, diabetes, BMI, waist-to-hip ratio, and systolic blood pressure, using genetic variants obtained from MR-base as above. Potential confounders that had significant (p<0.05) direct effects on atrial fibrillation in the individual multivariable models were then combined in a final model to jointly estimate their direct effects on atrial fibrillation.

Bidirectional MR was performed to assess for the possibility of reverse causality. A genetic instrument for atrial fibrillation containing 73 independent, genome-wide significant variants was constructed using the same methods as for the height instrument. Inverse-variance weighted, weighted median, and MR-Egger analyses were performed.

Individual-level MR was performed using the logistic control function estimator for the 6337 individuals with available height, genotype, and clinical covariate information.[30] The instrumental variable was a standardized genetic risk score for height, computed from independent, genome-wide significant variants, weighted by effect on height in the GIANT GWAS (see Height Genetic Risk Score above). Of the 707 genetic variants included in the main instrumental variable for height in the two-sample MR analysis, 695 were available for use in the individual-level analysis (**Supplemental Table 3**). In the first stage of the two-stage process, a linear regression was conducted with standardized height as the dependent variable, and the standardized genetic risk score for height as the independent variable. In the second stage, a logistic regression model with robust standard errors was fit, incorporating both the residuals from the first stage and scaled height, with atrial fibrillation as the outcome. In additional models, the second stage was adjusted for clinical diagnoses and left atrial size. Sensitivity analysis was performed including only individuals who had available echocardiographic data.

### Penn Medicine Biobank

The Penn Medicine Biobank is a longitudinal genomics and precision medicine study in which participants consent to linkage of genomic information and biospecimens to the electronic health record. More than 60,000 individuals are currently enrolled. This study included 7,023 individuals of European ancestry (genetically determined) who underwent genotyping and had available electronic health record data. For individual-level analyses in the Penn Medicine Biobank, continuous demographic variables were summarized by median and interquartile range, with categorical variable summarized by count and group-percent. Wilcoxon rank sum and Fisher’s exact tests, respectively, were used to assess for significant differences between groups.

### Phenotype Ascertainment

For individual level analyses in Penn Medicine Biobank, phenotypes were determined by querying the electronic health record. International Classification of Diseases (ICD) 9/10 and Current Procedural Terminology (CPT) codes, in addition to laboratory measurements and vital signs, were used to identify height, weight, BMI, smoking status, diagnoses of heart failure, hypertension, diabetes mellitus, chronic kidney disease, sleep apnea, stroke, thyroid disease, valvular heart disease, and cardiac surgery. Of the 7,420 individuals with high quality genotype data, complete phenotypes could be determined for 7,023 individuals. Transthoracic echocardiogram data was available for 2805 individuals. Left atrial size measured in the parasternal long axis view was extracted, and for individuals with more than one measure available the median value was used. Data was extracted as of January 2017. **(Supplemental Methods)**.

### Height Genetic Risk Score

A standardized, weighted genetic risk score (GRS) for height was calculated for each individual in the Penn Medicine Biobank using imputed dosage information in an additive model, weighted by the effect size of each independent (distance threshold = 10,000kb, r^2^ = 0.001), genome-wide significant (p < 5×10-8) variant from the Yengo et al. (2018) GWAS of height, using the *clump_data* command in the *TwoSampleMR* package in R.[19] Scores were standardized, centered to mean = 0 and scaled.

### Phenome-wide association study

Phenome-wide association studies were performed to identify clinical diagnoses associated both measured height (N = 6759) and the height genetic risk score (N = 7023), using the default setting of the *PheWAS* package in R.[31] Briefly, international classification disease codes obtained from the electronic health record were mapped to “Phecodes,” and individuals were assigned to case/control status or excluded using default mapping parameters. Association between height/height GRS and phenotypes was tested using logistic regression, adjusted for age, age^2^, sex, and 10 genetic principal components.

### Statistical Analysis

All statistical analyses were performed using R version 3.5.1.[32] For statistical analyses p<0.05 was considered statistically significant.

## RESULTS

### Demographics

Individual-level analysis focused on 7023 European Ancestry individuals from the Penn Medicine Biobank with available genotype data linked to the electronic health record.Individuals with atrial fibrillation were older (68 vs. 60 years; p < 0.001) than individuals without atrial fibrillation. Individuals with atrial fibrillation were taller (68 vs. 67 inches; p < 0.001), and significantly more likely to have been diagnosed with chronic kidney disease, heart failure, hyperlipidemia, hypertension, prior cardiac surgery, sleep apnea, smoking, stroke, thyroid disease, or valvular heart disease, compared to individuals without atrial fibrillation **(Table 1)**.

**Table 1:** Demographics.

|  | **No Atrial Fibrillation**<br>**(N = 3,803)** | **Atrial Fibrillation**<br>**(N = 3,220)** | **P-value** |
| --- | --- | --- | --- |
| **Age** |  |  |  |
| Median (IQR) | 61.0 (53.0, 68.0) | 65.0 (58.0, 74.0) | *P* < 0.001 |
| **Male** |  |  |  |
| n (%) | 2,110 (55) | 2,230 (69) | *P* < 0.001 |
| **BMI (kg/m<sup>2</sup>)** |  |  |  |
| n; Median (IQR) | 3,598; 28.0 (24.0, 32.0) | 3,180; 28.0 (25.0, 32.0) | *P* < 0.001 |
| **Height (in)** |  |  |  |
| n; Median (IQR) | 3,598; 67.0 (64.0, 70.0) | 3,180; 69.0 (65.0, 71.5) | *P* < 0.001 |
| **Weight (lbs)** |  |  |  |
| n; Median (IQR) | 3,598; 179.0 (150.0, 210.0) | 3,180; 189.5 (159.0, 222.0) | *P* < 0.001 |
| **Hypertension** |  |  |  |
| n (%) | 1,937 (51) | 2,244 (70) | *P* < 0.001 |
| **Coronary Artery Disease** |  |  |  |
| n (%) | 1,884 (50) | 1,902 (59) | *P* < 0.001 |
| **Heart Failure** |  |  |  |
| n (%) | 1,032 (27) | 1,799 (56) | *P* < 0.001 |
| **Hyperlipidemia** |  |  |  |
| n (%) | 1,991 (52) | 2,078 (65) | *P* < 0.001 |
| **Diabetes** |  |  |  |
| n (%) | 644 (17) | 534 (17) | *P* = 0.701 |
| **Chronic Kidney Disease** |  |  |  |
| n (%) | 434 (11) | 622 (19) | *P* < 0.001 |
| **Sleep Apnea** |  |  |  |
| n (%) | 441 (12) | 666 (21) | *P* < 0.001 |
| **Stroke** |  |  |  |
| n (%) | 615 (16) | 769 (24) | *P* < 0.001 |
| **Hyperthyroidism** |  |  |  |
| n (%) | 73 (2) | 86 (3) | *P* = 0.036 |
| **Cardiac Surgery** |  |  |  |
| n (%) | 881 (23) | 2,180 (68) | *P* < 0.001 |
| **Valve Disease** |  |  |  |
| n (%) | 978 (26) | 1,508 (47) | *P* < 0.001 |
| **Smoking (Ever)** |  |  |  |
| n (%) | 1,765/3,319 (53) | 1,621/2,899 (56) | *P* < 0.001 |

Demographics of individuals in Penn Medicine Biobank cohort with and without atrial fibrillation. kg = kilograms, m = meters, in = inches, lbs = pounds.

### Phenome-wide association studies of Height and Height GRS

To determine the association between genetically-predicted height and clinical diagnoses, we first constructed a genetic risk score for height using weights derived from the 2018 UK Biobank/GIANT height GWAS meta-analysis. Phenome-wide association studies were performed to identify clinical diagnoses associated with both measured height and the height genetic risk score across the Penn Medicine Biobank. Each 1 standard deviation increase in measured height was associated with increased risk of atrial fibrillation (OR 1.1; p = 5.9×10^−17^), and decreased risk of coronary atherosclerosis (OR 0.90; p = 3.4×10^−20^) and ischemic heart disease (OR 0.91; p = 1×10^−18^). Each 1 standard deviation increase in height PRS was associated with increased risk of atrial fibrillation and flutter (OR 1.15; p = 6×10^−6^) **(Figure 1)**.

**FIGURE 1:**
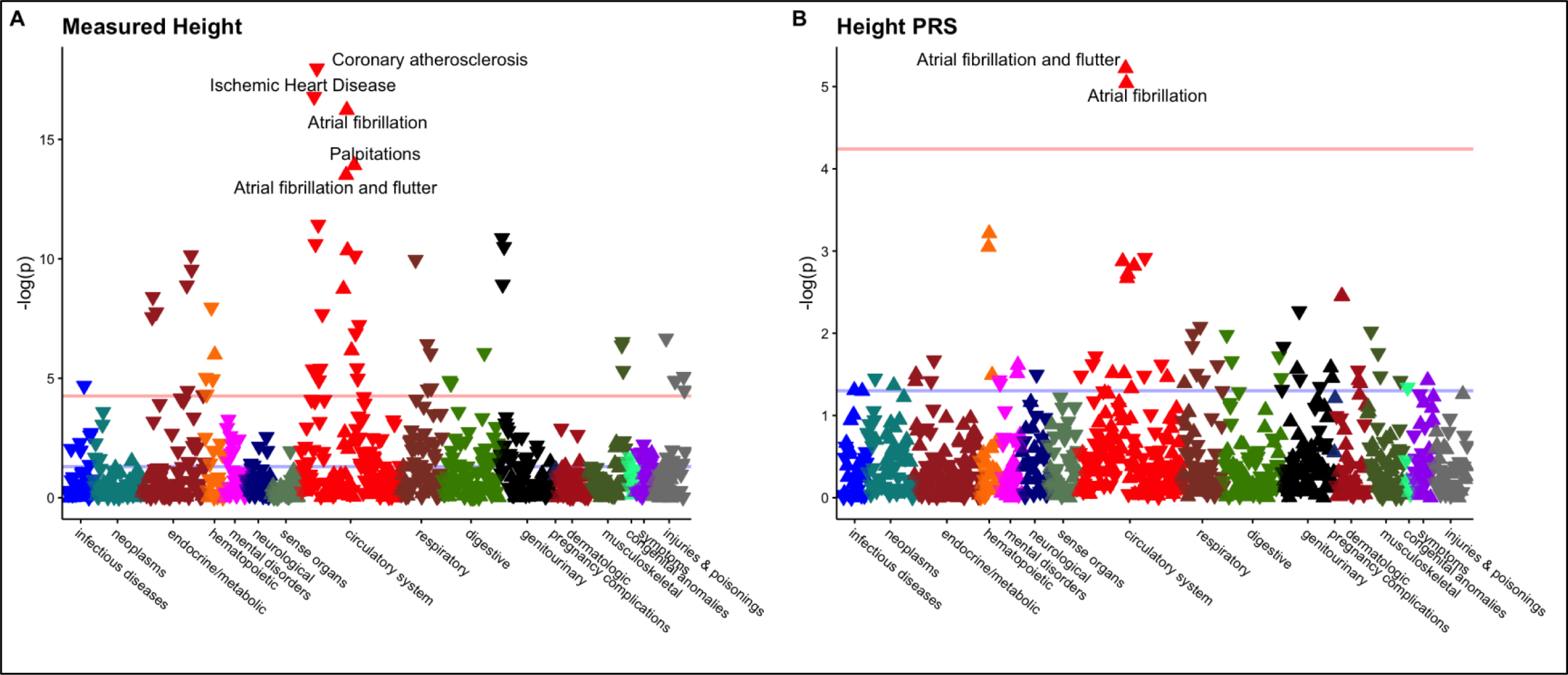
Phenome-Wide Associations with Height and Height GRS. Phenome-wide association studies were performed in Penn Medicine Biobank participants to identify clinical phenotypes associated with increased (A) height and (B) height GRS. The horizontal red line denotes the Bonferroni-adjusted level of significance (0.05/1816 phenotypes), and triangles denote the direction of association between increasing height and risk of the phenotype (point upward = increased risk; point downward = decreased risk).

### Population-Level Mendelian Randomization

To further characterize the relationship between increasing height and risk of atrial fibrillation, we constructed a genetic instrument for height using 707 independent SNPs associated with height at a genome-wide level of significance (p < 5×10^−8^), which accounted for 11.2% of the variability in height **(Supplemental Table 1)**. The mean F-statistic was 110 (range 22-948), suggesting the risk of weak-instrument bias was low.[33]. We performed two-sample Mendelian randomization using summary statistics from a genome-wide association study of atrial fibrillation including 65,446 atrial fibrillation cases and 522,744 controls. Inverse variance-weighted modeling identified a significant association between increasing height and atrial fibrillation (OR 1.34; 95% CI 1.29 to 1.40; p = 5×10^−42^) **(Figure 2)**. The intercept from Egger regression was -0.001 (p = 0.13), thus not providing evidence for significant pleiotropic bias.Similar estimates were obtained in sensitivity analyses from weighted median, MR-Egger, and MR-PRESSO models.

**FIGURE 2:**
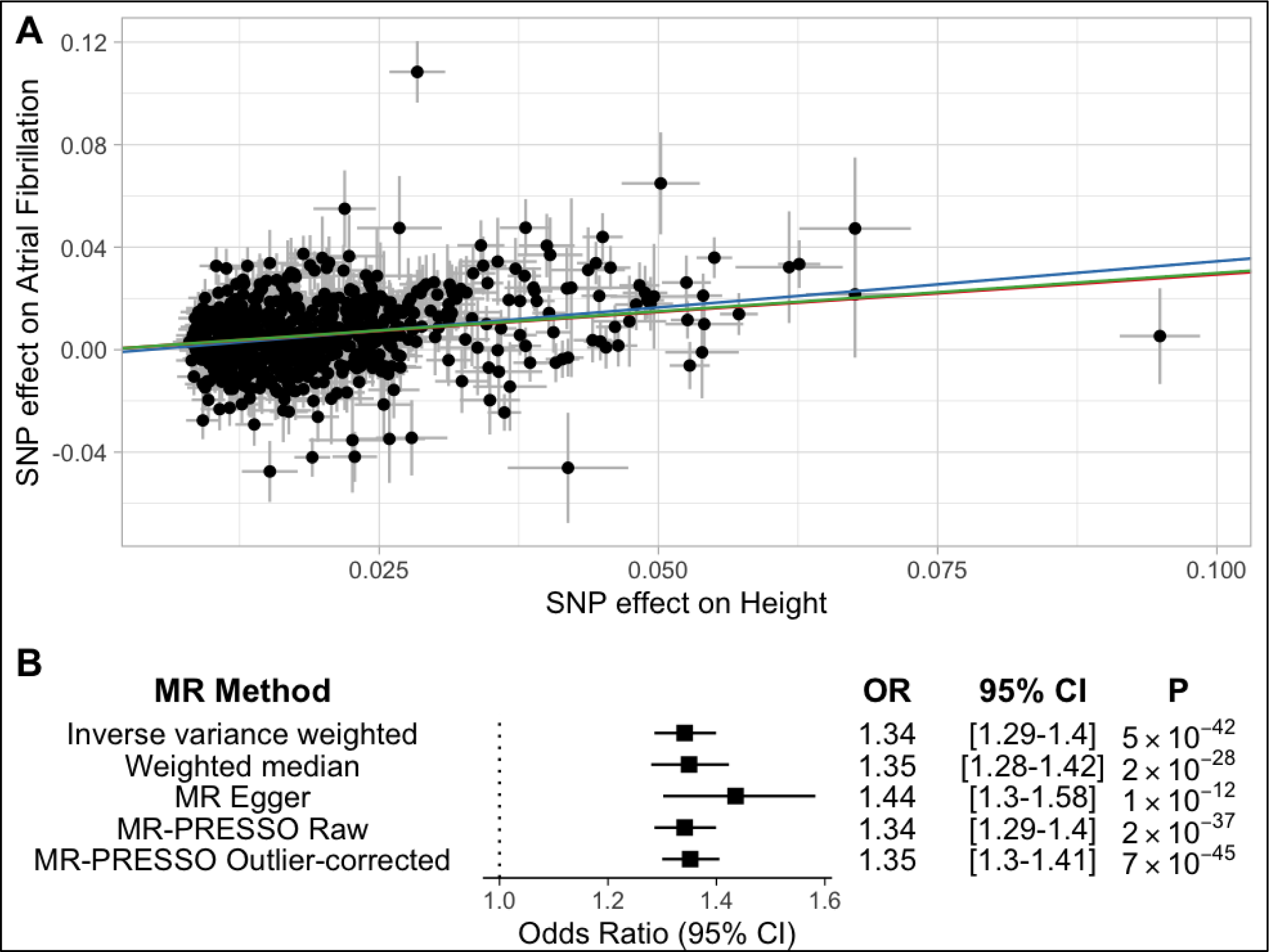
Two-Sample Mendelian Randomization. Two-sample Mendelian randomization was performed using a genetic instrument containing 707 independent SNPs associated with height. A) Each point represents the SNP effects on height and atrial fibrillation. Colored lines represent inverse variance weighted (red), weighted median (green), and MR-Egger (blue) estimates of the association between a 1 standard deviation increase in height and risk of atrial fibrillation. B) Odds Ratios (OR), 95% Confidence Intervals (CI) and P-values for Mendelian randomization estimates.

An additional genetic instrument for height was constructed, excluding variants nominally associated (p < 0.05) with potentially pleiotropic risk factors for atrial fibrillation. The restrictive instrument consisted of 224 independent SNPs, in sum explaining 2.8% of the variance in height **(Supplemental Table 2)**. The mean F-statistic was 88 (range 22-867). Mendelian randomization results using this restrictive genetic instrument were similar **(Supplemental Figure 1)**.

We next performed multivariable Mendelian randomization. Height remained significantly associated with atrial fibrillation after adjustment for the effect of genetic variants separately on each of coronary artery disease, HDL, LDL, total cholesterol, triglycerides, fasting glucose, fasting insulin, diabetes, BMI, waist-to-hip ratio, and systolic blood pressure **(Supplemental Table 4)**. This analysis identified significant associations between BMI, systolic blood pressure, total cholesterol, and coronary artery disease with atrial fibrillation after adjustment for height. The effect of height on risk of atrial fibrillation was similar in models accounting for these risk factors individually, and in a combined model jointly considering genetic variants associated with height, BMI, systolic blood pressure, total cholesterol, and coronary artery disease **(Figure 3)**.

**FIGURE 3:**
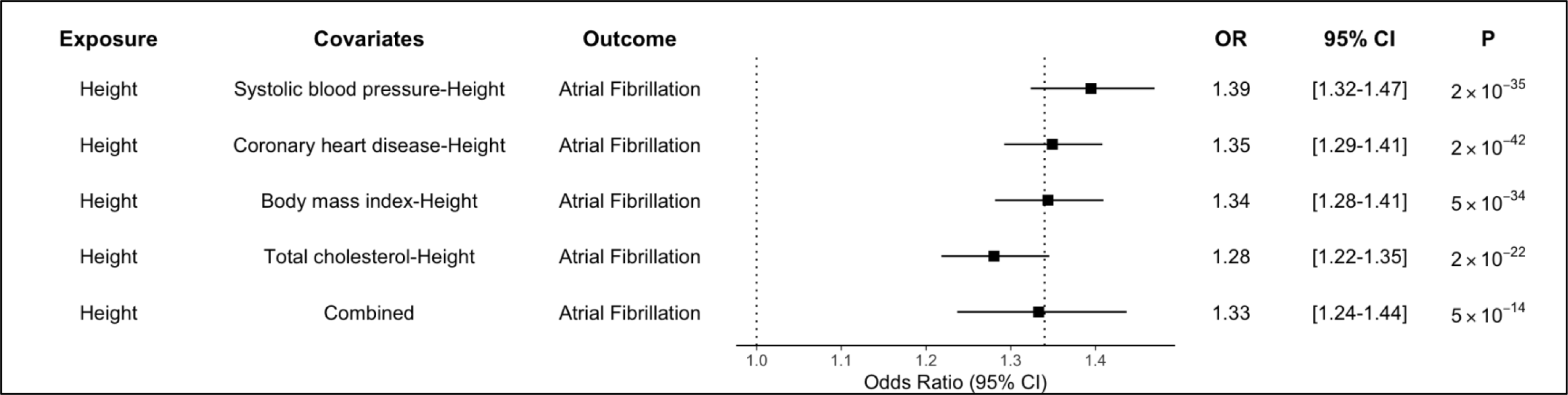
Multivariable Mendelian Randomization Analysis. Multivariable Mendelian randomization was performed to jointly consider the effect of genetic variants for cardiometabolic traits and height on atrial fibrillation. The dashed line at OR 1.34 represents the effect of height on atrial fibrillation observed in the primary inverse variance weighted analysis. Odds Ratios (OR), 95% Confidence Intervals (CI) and P-values for Mendelian randomization estimates are displayed.

To assess for the possibility of reverse-causation, where genetic variants primarily associated with increased risk of atrial fibrillation may represent invalid instruments for height, a bi-directional multivariable analysis was performed. Using a genetic instrument including 73 independent, genome-wide significant variants associated with atrial fibrillation, there was no evidence for a causal association with height **(Supplemental Figure 2)**.

### Individual-Level Mendelian Randomization

Individual-level Mendelian randomization was performed in Penn Medicine Biobank participants to further assess the association between height and atrial fibrillation. Using the height weighted genetic risk score as an instrumental variable, height was significantly associated with atrial fibrillation (OR 1.69 per 1 standard deviation increase in height; 95% CI 1.54 to 2.19; p = 5×10^−4^) after adjusting for age, sex, and 5 genetic principal components **(Figure 4)**. Height remained associated with atrial fibrillation after adjustment for weight, hypertension, coronary artery disease, heart failure, hyperlipidemia, diabetes, chronic kidney disease, sleep apnea, stroke, hyperthyroidism, smoking, cardiac surgery, and valvular heart disease. After further adjustment for left atrial size, height remained significantly associated with atrial fibrillation (OR 1.76; 95% CI 1.43 to 2.17; p = 0.005). When individual-level MR was restricted to the subset of individuals with complete data, the effect estimates were similar, with modest attenuation with sequential adjustment for clinical risk factors and left atrial size **(Supplemental Figure 3)**.

**FIGURE 4:**
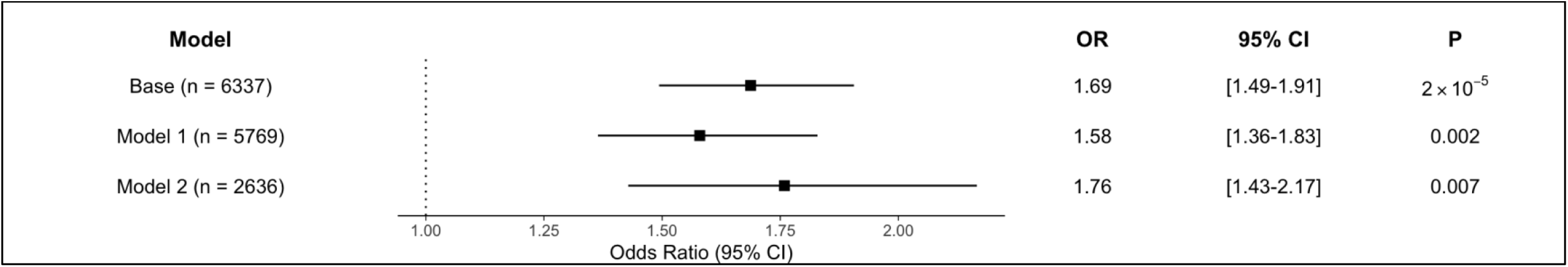
Individual-Level Instrumental Variable Analysis. Individual-level instrumental variable analysis was performed in Penn Medicine Biobank participants, using a weighted genetic risk score for height as an instrumental variable for measured height. The Base model was adjusted for age, sex, and 5 genetic principal components. Model 1 additionally adjusted for weight, hypertension, coronary artery disease, heart failure, hyperlipidemia, diabetes, chronic kidney disease, sleep apnea, stroke, hyperthyroidism, smoking, cardiac surgery, and valvular heart disease. Model 2 additionally adjusted for left atrial size as measured on transthoracic echocardiogram. Odds ratio are reported per 1 standard deviation increase in height.

## DISCUSSION

In this study we used both population- and individual-level genetic information to test the association between height and atrial fibrillation. At the population level, there was a strong causal association between genetic determinants of height and risk of atrial fibrillation. This finding was robust to multiple sensitivity analysis in the Mendelian randomization methods and the genetic instrument for height. Observational analysis at the individual-level identified a strong association between height and increased risk of atrial fibrillation, and decreased risk of coronary artery disease. Using individual-level MR analysis, we found that the relationship between height and atrial fibrillation appears to be independent from traditional clinical and echocardiographic risk factors for atrial fibrillation.

Our findings are consistent with prior observational analyses that have identified height as a risk factor for atrial fibrillation.[4,11–18] These studies, including a large Swedish national cohort study of 1.5 million military conscripts recruited over the course of 28 years have consistently identified a strong association between height and atrial fibrillation.[4] Observational designs have limited these studies, due to the possibility of residual confounding. The Helsinki Birth Cohort Study partially addressed this limitation by considering the effect of maternal height on risk of atrial fibrillation in offspring, but was limited by a small, homogenous sample.[34] We further build on those prior findings using the Mendelian randomization framework, considering both summary-level and individual-level genetic data, leveraging genetic variants associated with height and atrial fibrillation to provide a causal estimate for the effect of increasing height on risk of atrial fibrillation. Our population-level Mendelian randomization analysis reinforces a prior MR estimate for association between increasing height and atrial fibrillation (OR 1.33; 95% CI 1.26 to 1.40) using updated GWAS summary statistics for height and atrial fibrillation.[9] By leveraging large multiethnic studies of both height and atrial fibrillation, we increase the generalizability of the prior MR findings, which were limited only to white participants of the UK Biobank.

Several mechanisms have been proposed to explain the relationship between height and atrial fibrillation. Increased left atrial size is correlated with height, and has been identified as independent predictor of atrial fibrillation.[18,35–37] Consistent with findings from the Cardiovascular Health Study, however, we found the effect of height on risk of atrial fibrillation was not attenuated after adjustment for left atrial diameter.[14] It is possible that other, more nuanced markers of left atrial structure and function that have been associated with severity of atrial fibrillation, such as left atrial volume index, emptying fraction, expansion index, and contractile function, may also be affected by height and may better explain the association between height and atrial fibrillation.[38] Similarly, two-dimensional echocardiography significantly underestimates left atrial size compared to MRI assessment.[39] Investigation of these factors in the two-sample setting is limited by the lack of genetic studies of cardiac structure/function by echocardiography, MRI, or cardiac CT, and study of these parameters in large cohorts with genetics and imaging data is warranted.

Mendelian randomization has previously identified an association between shorter stature and increased risk of coronary artery disease, mediated in part by increases in LDL cholesterol and triglyceride levels.[10] In the current analysis, we were unable to detect significant associations between these lipid traits and risk of atrial fibrillation when considered in multivariable models alongside height. Similarly, in multivariable models considering height alongside common cardiometabolic risk factors for atrial fibrillation, effect estimates were all similar, suggesting these factors may not substantially mediate the effect of height on risk of atrial fibrillation.

Although we detected no evidence of horizontal pleiotropy and our results remained robust to extensive sensitivity analyses, height is a highly polygenic trait, and the possibility remains that pleiotropic pathways mediate the association between height and atrial fibrillation. A recent genome wide association study found genes at atrial fibrillation-associated loci to be enriched in pathways important for tissue formation.[40] Coupled with the finding that genes at height-associated loci are enriched for cardiovascular and endocrine tissue types, these results raise the possibility of a more complex shared genetic architecture affecting both height and atrial fibrillation.[22] Thus, we cannot exclude the possibility that genetic variants broadly associated with growth and development may simultaneously affect height and establish structural cardiovascular changes that may predispose to atrial fibrillation.

Our study has several limitations. First, observational analyses from the Penn Medicine Biobank may not be generalizable, as this population represents a cohort within a single academic health system. Second, despite our use of multiethnic GWAS of height and atrial fibrillation, the underlying studies focused primarily on individuals of European ancestry. Genetic studies in broader populations are warranted to further improve generalizability of these findings. Third, our population-level MR analyses relied on publicly available summary statistics which contain some overlapping samples/cohorts. Two-sample MR tends to bias the causal effect estimates to the null, but sample overlap may make the estimate susceptible to weak-instrument bias. In this study, however, despite overlapping cohorts, simulation suggests the large sample sizes of the height and atrial fibrillation GWAS, and large F-statistics make the risk of weak instrument bias low.[33]

In conclusion, we find that increases in height are associated with increased risk of atrial fibrillation, and this relationship is likely to be causal. These results raise the possibility of investigating height/growth-related pathways as a means for gaining novel mechanistic insights to atrial fibrillation, as well as the possibility of incorporating height into larger targeted screening strategies for atrial fibrillation.

## Data Availability

GWAS summary statistics for height GWAS can be downloaded from the GIANT consortium website (https://portals.broadinstitute.org/collaboration/giant/index.php/GIANT_consortium_data_files). Summary statistics for atrial fibrillation were contributed by the AFGen consortium (http://afgen.org) and downloaded from the Variant to Function Knowledge Portal (http://www.kp4cd.org/datasets/v2f). The data, analytic methods, and study materials may be made available to other researchers for purposes of reproducing the results or replicating the procedure upon reasonable request to the corresponding author and with the appropriate ethical approval and data sharing agreements.

## ACKNOWLEDGEMENTS

The authors would like to thank the participants of the Penn Medicine Biobank.

## ETHICAL APPROVAL

The Penn Medicine Biobank has received approval from the University of Pennsylvania Institutional Review Board. No additional approval was required for analyses of publicly available summary statistics.

## TRANSPARENCY

The lead author (MGL) affirms that the manuscript is an honest, accurate, and transparent account of the study being reported; that no important aspects of the study have been omitted; and that any discrepancies from the study as originally planned (and, if relevant, registered) have been explained.

